# Estimating the Burden of SARS-CoV-2 among the Rohingya Refugees

**DOI:** 10.1101/2021.06.11.21258445

**Authors:** Shaun A. Truelove, Sonia Hegde, Lori Niehaus, Natalya Kostandova, Chiara Altare, V. Bhargavi Rao, Julianna Smith, Philipp du Cros, Andrew S. Azman, Paul Spiegel

## Abstract

**Background:** Since the emergence of the COVID-19 pandemic, substantial concern has surrounded its impact among the Rohingya refugees living in the Kutupalong-Balukhali refugee camps in Bangladesh. Early modeling work projected a massive outbreak was likely after an introduction of the SARS-CoV-2 virus into the camps. Despite this, only 317 laboratory-confirmed cases and 10 deaths were reported through October 2020. While these official numbers portray a situation where the virus has been largely controlled, other sources contradict this, suggesting the low reported numbers to be a result of limited care seeking and testing, highlighting a population not willing to seek care or be tested. SARS-CoV-2 seroprevalence estimates from similar a timeframe in India (57%) and Bangladesh (74%) further sow doubt that transmission had been controlled. Here we explore multiple data sources to understand the plausibility of a much larger SARS-CoV-2 outbreak among the Rohingya refugees.

**Methods:** We used a mixed approach to analyze SARS-CoV-2 transmission using multiple available datasets. Using data from reported testing, cases, and deaths from the World Health Organization (WHO) and from WHO’s Emergency Warning, Alert, and Response System, we characterized the probabilities of care seeking, testing, and being positive if tested. Unofficial death data, including reported pre-death symptoms, come from a community-based mortality survey conducted by the International Organization for Migration (IOM),) in addition to community health worker reported deaths. We developed a probabilistic inference framework, drawing on these data sources, to explore three scenarios of what might have happened among the Rohingya refugees.

**Results:** Among the 144 survey-identified deaths, 48 were consistent with suspected COVID-19. These deaths were consistent with viral exposures during Ramadan, a period of increased social contacts, and coincided with a spike in reported cases and testing positivity in June 2020. The age profile of suspected COVID-19 deaths mirrored that expected. Through the probability framework, we find that under each scenario, a substantial outbreak likely occurred, though the cumulative size and timing vary considerably. In conjunction with the reported and suspected deaths, the data suggest a large outbreak could have occurred early during spring 2020. Furthermore, while many mild and asymptomatic infections likely occurred, death data analyzed suggest there may have been significant unreported mortality.

**Conclusions:** With the high population density, inability to home isolate adequately, and limited personal protective equipment, infection prevention and control in the Rohingya population is extremely challenging. Despite the low reported numbers of cases and deaths, our results suggest an early large-scale outbreak is consistent with multiple sources of data, particularly when accounting for limited care seeking behavior and low infection severity among this young population. While the currently available data do not allow us to estimate the precise incidence, these results indicate substantial unrecognized SARS-CoV-2 transmission may have occurred in these camps. However, until serological testing provides more conclusive evidence, we are only able to speculate about the extent of transmission among the Rohingya.

## Background

Soon after the emerging COVID-19 epidemic had been declared a pandemic, public health authorities, researchers, and humanitarian organizations began mobilizing efforts to address this imminent threat in one of the most vulnerable and largest refugee populations in the world, the Rohingya living in Cox’s Bazar, Bangladesh. Early estimates projected substantial impact in this population, with hundreds of thousands of infections leading to 20,000-30,000 hospitalizations and upwards of 2,000 deaths – in a population with extremely limited capacity.^1^ Humanitarians and health officials quickly agreed that control measures like physical distancing, home isolation, and handwashing would be extremely challenging.^2–4^ As a result, isolation treatment centers were rapidly built, border control was quickly established, and mass gatherings were canceled.^5–7^

On 14 May 2020, the first cases of COVID-19 were confirmed among the Rohingya refugees, and the humanitarian community braced for the worst.^8^ Exactly one year after the first case was detected, as of 14 May 2021 only 690 cases and 11 deaths had been confirmed among this population.^9^ If the scale of the reported case and death numbers is correct, this would likely represent a major success of the control measures put in place to stop viral introductions and slow transmission. However, several notable contradictions to this narrative persist. First, while control measures may have limited mass gatherings, movement in and out of camps, and produced hundreds of beds in isolation centers, the effectiveness of these measures to limit transmission relies on rapidly capturing and isolating a high proportion of cases and contacts.^10^ The continued positive cases identified in this population alone indicate transmission has continued at some level, particularly if infection importation has been controlled.

Serology has begun to shed light on challenges like these, and numerous examples exist of serological surveys demonstrating extensive transmission in populations believed to have been spared. In the Indian slum Dharavi in Mumbai, a serological survey in July 2020 found 57% seroprevalence though only a month prior it had been touted as a global example of success.^11,12^ Other slums in India, Bangladesh, and Argentina demonstrated similar findings in populations with otherwise small numbers of reported cases.^13–16^ In each example, confirmed cases captured an insignificant fraction of what the serology implied, and confirmed deaths were limited.

To overcome limited detection of mild or asymptomatic infections, the ratio of reported cases to infections has been used to estimate the total number of SARS-CoV-2 infections.^17^ However, reports of widespread influenza-like-illness, unconfirmed deaths, avoidance of health care, and unwillingness to be tested in Cox’s Bazar refugee camps highlight a context where extrapolation from the reported burden using a constant ratio may mischaracterize the true burden.^18–21^ Challenges around barriers and willingness to seek care and be tested for SARS-CoV-2 infection have not been unique to the Rohingya during this pandemic. However, these challenges are likely magnified in this population. This lack of willingness to report illness and death may stem from a long history of mistrust, persecution, and violence against the Rohingya, mistreatment that eventually culminated in a mass exodus of over 700,000 to Bangladesh to escape persecution, some claiming genocide, in their home country of Myanmar in 2017.^22^

In this analysis, we assess available sources of data on SARS-CoV-2 and COVID-19 among the Rohingya refugees, including counts of confirmed cases and deaths, polymerase chain reaction (PCR) testing data, and survey data on unconfirmed deaths. We explore what these data indicate in terms of transmission and disease burden, where these data disagree, and where they align. We explore support for three plausible scenarios where:

*Scenario 1: transmission was relatively limited, with infections generally proportional to reported cases;*

*Scenario 2: a limited early outbreak occurred that is not captured by the official case counts; and*

*Scenario 3: a large, widespread outbreak occurred, and transmission was largely missed*.

Through these analyses, we aim to better understand what these data sources collectively indicate and the degree to which they provide support for each of the three scenarios.

## Methods

The purpose of this study was to assess possible scenarios of SARS-CoV-2 transmission that may have occurred among the Rohingya refugees living in 34 refugee camps that constitute the Kutupalong-Balukhali refugee camps, located in Cox’s Bazar District, Bangladesh, comprising a total population of 866,457.^23^ We approached this analysis through two distinct, complementary analyses. First, we analyzed deaths from a mortality survey to classify those from suspected COVID-19, from which we inferred the number of infections. Second, we developed a model to infer infections and deaths from official case and testing data, while accounting for additional data on care seeking and testing access over time.

To explore how changes in care-seeking behavior and willingness to be tested for SARS-CoV-2 infection could influence how many infections the official laboratory counts represent, we developed a probabilistic model to infer infection from testing data. This inference model allows us to explore how potential changes in care seeking and willingness to get tested, and testing availability independently change how should interpret what official positive cases represent, accounting for variations in positivity rates and absolute numbers tested and positive for SARS-CoV-2 infection.

### Data sources

The primary data sources used for this analysis included:

#### 1) Reported COVID-19 cases, deaths, and testing data

All data on confirmed cases and deaths and testing were acquired from the World Health Organization (WHO) through the WHO Cox’s Bazar Data Hub’s COVID-19 Dashboard.^9^ These data include weekly counts of PCR tests performed and weekly counts of PCR-positive cases and deaths by camp. These data cover the period 29 March to 31 October 2020 and are publicly available.

#### 2) Weekly acute respiratory infections (ARI)

We extracted weekly numbers of individuals seeking care for ARI from Early Warning, Alert, and Response System (EWARS) reports from January 2018 to December 2020.^24^ These were used to estimate the proportion of individuals with illness similar to COVID-19 that seek care.

#### 3) Suspected COVID-19 deaths

Unreported, suspected COVID-19 deaths were identified through a mortality survey conducted by the International Organization for Migration (IOM).^18^ A team of Rohingya researchers conducted the survey among the Rohingya population from 30 June to 11 July 2020 to capture unreported deaths that occurred between 22 March and 11 July 2020.^18^ The refugee population lives in 34 refugee camps, which includes a “megacamp” complex of 26 camps and 8 camps located further south. Each camp is subdivided into sub-blocks consisting around 100 households, each with a sub-block community leader, or “*mazi*”. The *mazis* are often responsible for any religious preparations or support provided to the family of the deceased, and thus have strong knowledge of deaths occurring in their sub-block. Three hundred fifty (18%) of the total 1,949 sub-blocks were randomly selected (population-weighted) and their *mazis* approached for interviews, 300 of whom (86%) consented to participate, representing 129,042 individuals from 30,866 households, and 15% of the total population of 866,457. Sub-blocks surveyed represented 24 of the 34 Kutupalong-Balukhali refugee camps. All sub-blocks from Camp 6 and the Kutupalong Refugee Camp (KRC) refused consent, and the 8 camps (Camps 21-27, Nayapara RC) located south of the “megacamp” were not included for logistical reasons. All data on deaths were provided retrospectively and are subject to issues of recall and willingness to provide information. For each death, the following data were recorded: age, sex, location, symptoms, other reported comorbid conditions, and cause of death, though clinical information was largely incomplete. We used the available symptoms, comorbid conditions, and cause of death to classify each of these deaths according to the case definitions reported below. We estimated the sampling-adjusted number of deaths at the camp-level and overall, by extrapolating from the proportion of individuals represented by the survey.

#### 4) Age-specific population by camp

We estimated the expected infection fatality ratio (IFR) using published 5-year age- and sex-specific probabilities of mortality following infection and refugee camp sub-block population data from United Nations High Commissioner for Refugees (UNHCR) (*see Supplemental Materials*).^25^ Sub-block-level IFR estimates were aggregated to get camp-specific and overall IFR adjusted for sub-block population, age, and sex.

All PCR-positive official deaths due to COVID-19 were checked against the mortality survey deaths, using camp, sex, and ranges for both age (+/-5 years) and week reported (+/-1 week). One potential match was found (a male reported from Camp 10 during week 24 with a 3-year difference in age [70 vs 67 years]); because of the similarity, these were considered the same individual.

### Survey case definitions

While some of the deaths captured by the mortality survey may be due to COVID-19, it is probable that some are not. For this analysis, we apply a set of case definitions to classify the deaths from the mortality survey (see *Supplementary Materials 2*.*1* for full details). We classified a “suspected COVID-19” case following the official U.S. Centers for Disease Control and Prevention (CDC) case definitions. Because the CDC case definition was more aligned with the data available from the survey, we use it, rather than the WHO case definition, in this analysis. Individuals dying from other reported causes were defined as “other cause”. All deaths for which data were incomplete or the cause of death was undefined were classified as indeterminate.

### Infection and testing model

In this probability framework, we use Bayes’ Theorem to express the probability of infection in the population at a specific time-point as:

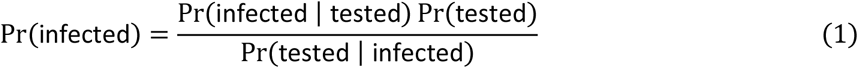

Here the *Pr* (infected) is the probability that any individual is infected during the specified time period; *Pr* (infected | tested) is the time-varying positivity rate, adjusting for test sensitivity (i.e., the probability that an individual tested is positive for SARS-CoV-2 infection); and *Pr* (tested) is calculated as the number of tests performed divided by the total population at each time-point (e.g., daily or weekly).

We further assume that to get tested in this population, an individual must seek care and be willing to be tested (“seek care & testing”), present with symptoms that meet the case definition for COVID-19 (“case definition”), and there must be testing availability at the time of care seeking. For this population and analysis, we assume testing of asymptomatic individuals is negligible. We represent the probability that an infected individual is tested in the population as

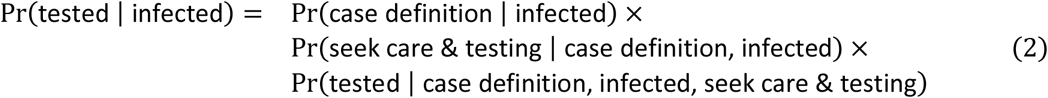

We estimate these probabilities using data available and as defined by three scenarios below.

### Scenarios

Individuals’ willingness to seek care and the availability of testing are not well known and can substantially shape our interpretation of reported case and death. We explored three different scenarios with varying assumptions about how people’s willingness to seek care for COVID-19-like illness and their ability to get a SARS-CoV-2 test changed over the course of the epidemic.

For each scenario, we estimate the expected weekly number of infections and deaths using the estimated age-adjusted IFR for the Rohingya population. We examine these resulting infections and deaths in comparison to the reported, confirmed COVID-19 deaths and suspected deaths, weighted and unweighted. These scenarios and their parameters are not meant to be exact, as the data to precisely inform them are not available. Instead, these are meant to explore three potential phenomenological perspectives of what occurred during the COVID-19 pandemic among the Rohingya, within the confines of the reported data (i.e., confirmed cases, deaths, and testing data). Full details of the model and how each set of data are used is detailed in the *Supplemental Materials 1*.*2*.

#### Scenario 1. Care seeking proportional to infection (null scenario)

This scenario serves as our status quo scenario, in which it is assumed care-seeking behavior did not change over time and reported PCR-positive cases are roughly proportional to the underlying infections in the population. This use of confirmed cases represents a common interpretation of the progression of the pandemic among the Rohingya and in most populations around the world. We explore this scenario assuming that *Pr*(case definition | infected)=0.10 and *Pr*(seek care & testing | case definition, infected)=0.22, based on data from a Dhaka seroprevalence study and the number of reported cases in Dhaka at that time (Figure 3A).^26,27^ We further assume that 60% of individuals that seek care and meet the testing criteria are tested.^20^ *See Supplementary Materials 1*.*2 for details about how these parameters have been estimated*.

#### Scenario 2. Time-varying care seeking

This scenario assumes a baseline, pre-pandemic probability of seeking care for ARI that is equal to that of Scenario 1. However, effective care-seeking is allowed to vary over time, defined as *Pr* (seek care & testing | case definition, infected)_*t*_ = *Pr*(seek care & testing | case definition, infected)_0_ * *αt*. We express *α*_*t*_ as the ratio of observed to expected ARI cases in the population using EWARS ARI data (*see Supplementary Materials 1*.*2*). Like Scenario 1, we assume *α*_*t*_ (case definition | infected) =0.10 and that 60% of individuals that seek care and meet the testing criteria are tested.

#### Scenario 3. Time-varying care seeking and testing

For the final scenario, we assume time-varying care-seeking behavior that follows the observed ARI cases as in Scenario 2, but also incorporate time-varying testing. We quantify *Pr*(tested | infected) empirically using the expected EWARS ARI counts, the number tested, and *Pr*(case definition | infected). We assume probabilities of seeking care and testing and of being tested are equivalent whether an individual is SARS-CoV-2 infected or not. This allows for the quantification of

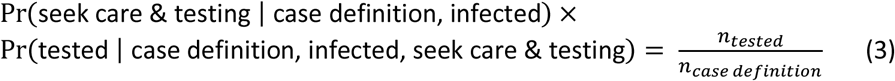

Applying this to Equation 2, we estimate a time-varying *Pr*(tested | infected) tested.

## Results

From 14 May to 31 October 2020, 317 laboratory-confirmed cases of COVID-19 were reported among the Rohingya in Cox’s Bazar, occurring in seemingly two waves (Figure 1A) with confirmed cases reported in each of the 34 camps (Figure 2C). Among these cases, ten died (Confirmed Case Fatality Risk=3.15%), including six deaths in May-July 2020 (Figure 2A).

**Figure 1.**
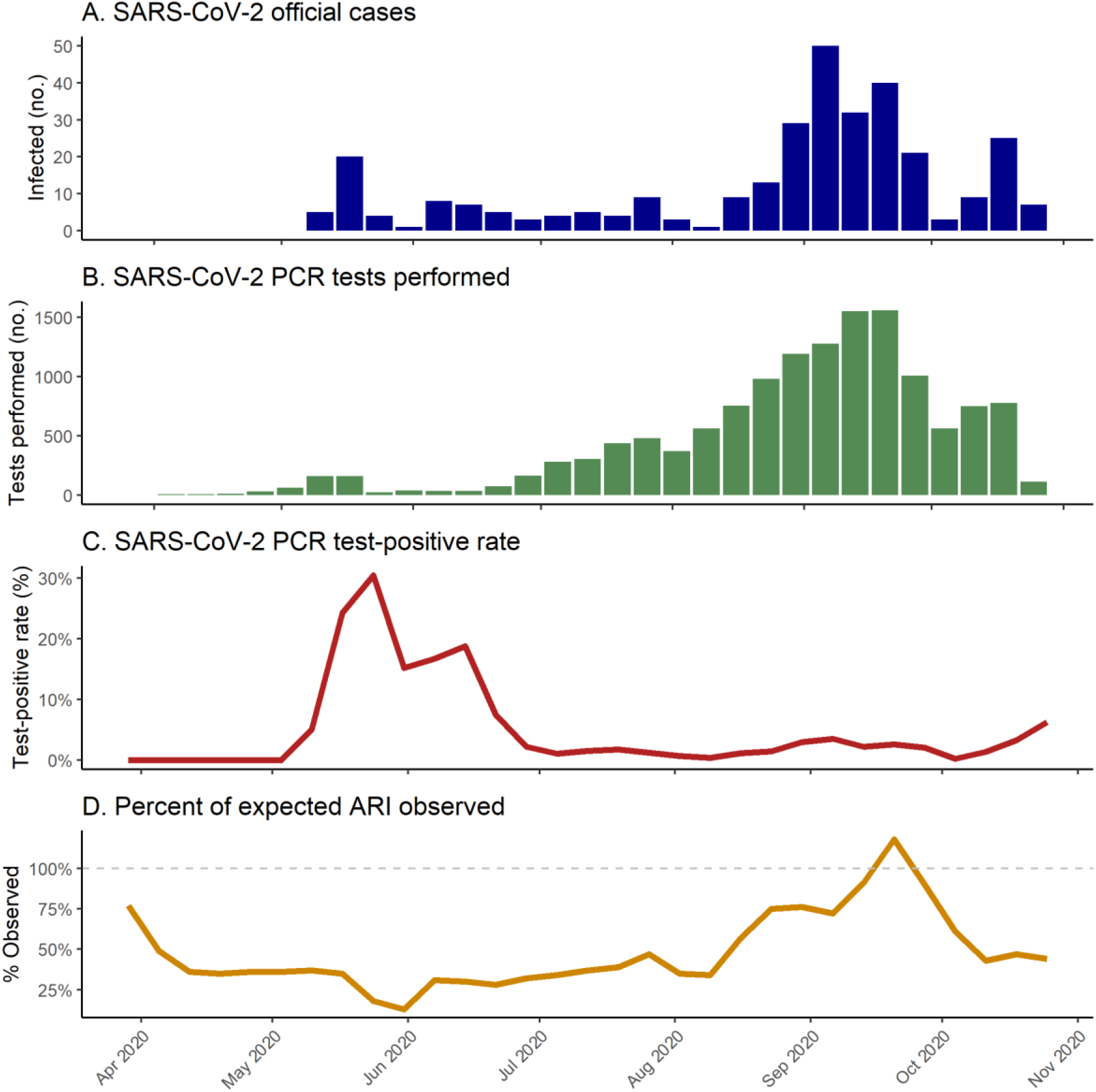
SARS-CoV-2 incident cases, number tested, test-positivity, and care seeking changes among the Rohingya refugees. The number of SARS-CoV-2 infections that are detected and confirmed though RT-PCR testing (A) is a function of multiple factors: the number of tests performed (B), the test-positivity rate (C), and the proportion of those infected who seek care and confirmatory testing, which may be represented by the percent of the estimated expected acute respiratory infection (ARI) that were observed (D). The test-positivity rate is often considered to reflect the amount of infection in the population, while the number of tests performed can be biased by both testing capacity and willingness to seek care and be tested. Willingness to seek care may be reflected in overall ARI surveillance data, such as those provided by the World Health Organization’s Emergency Warning, Alert and Response System (EWARS)^24^. Reductions in care seeking and reporting of ARI may point to reduced willingness to seek care among the Rohingya refugees during this pandemic period. Considered together, these data may provide a clearer picture of the true number of infections over time in the population, which remains unknown. Data on reported cases, tests performed, and test-positivity are from the WHO Cox’s Bazar Data Hub’s COVID-19 Dashboard (A-C)^9^. Estimates of percentage of expected ARI that are observed (D) were estimated as the number of ARI observed during 2020 divided by the number estimated by fitting a multilevel linear regression model to EWARS data from 2018-2019 (see *Supplementary Materials 1*.*2*).

**Figure 2.**
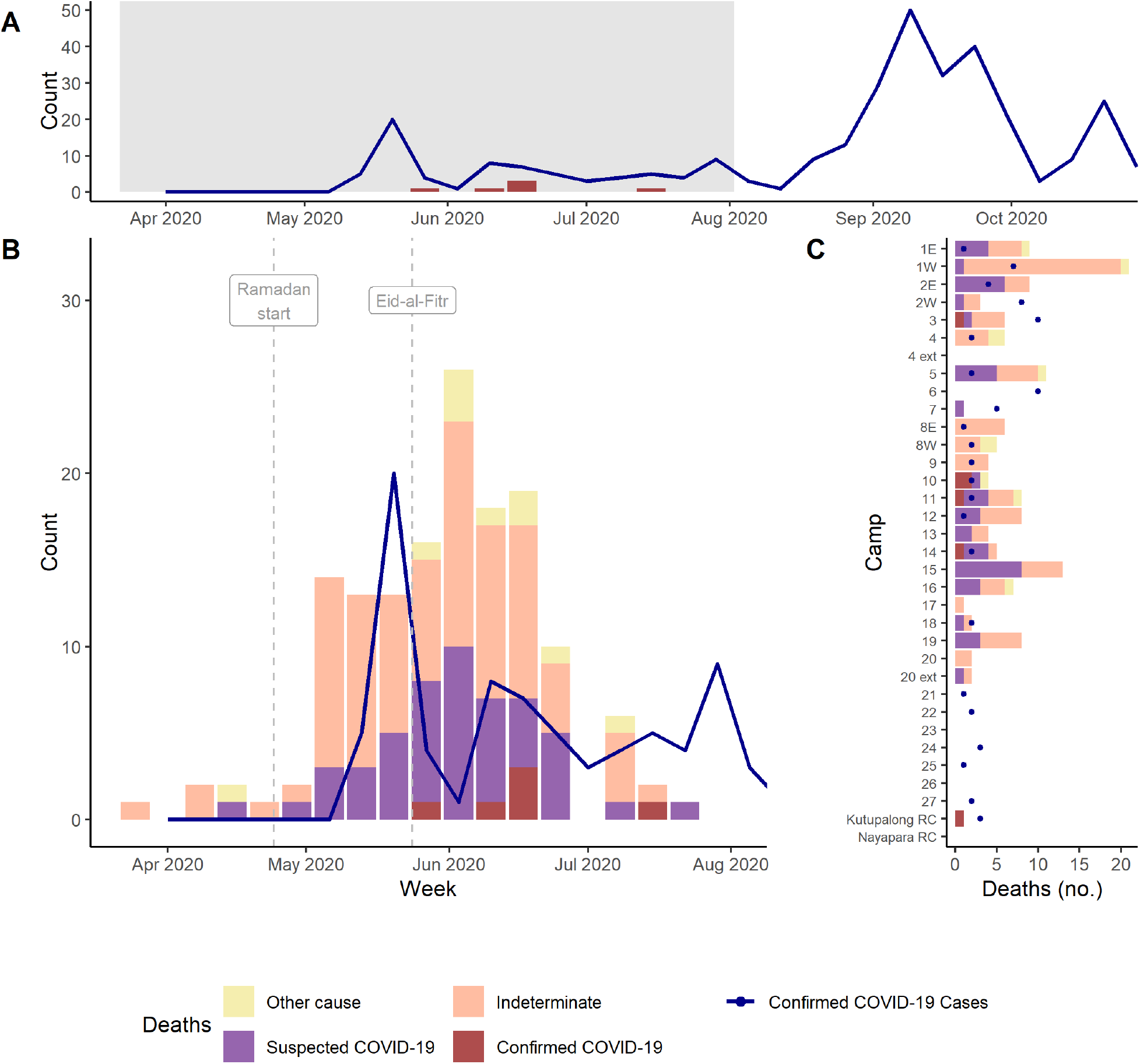
Confirmed COVID-19 cases and deaths and suspected COVID-19 deaths among the Rohingya Refugees in Cox’s Bazar, Bangladesh. (A) Official PCR-positive confirmed COVID-19 cases and deaths, May-October 2020. The grey box represents the time period from which deaths were identified by the survey conducted by the International Organization for Migration (IOM), which are represented in (B). (B) Deaths in the mortality survey by cause (yellow, pink, purple) and reported PCR-positive COVID-19-related deaths (maroon bars), by week of death, March-July 2020. Overlap with confirmed cases is displayed (navy blue line and points). (C) Camp of residence of PCR-positive deaths, unreported deaths, and reported confirmed PCR-positive COVID-19 cases in the Kutupalong-Balukhali refugee camps, Cox’s Bazar, Bangladesh, March-July 2020.

During this study period, a total of 13,783 SARS-CoV-2 PCR tests were performed among this population, with an overall positivity rate of 2.3% (Figure 1B). Positivity has varied substantially over time, with higher positivity observed at the start of testing, peaking at 26.4% in the first week of June 2020 and dropping to 1-6% during July-October 2020 (Figure 1C).

Care seeking for acute respiratory illness declined within the Rohingya population during the pandemic. A sharp drop in ARI was reported by community health workers to EWARS starting the week of 16 March 2020, declining to 13% of expected ARI levels the week of 31 May 2020, before increasing slowly towards expected levels (Supplementary Figure S1).

### Survey-identified deaths

The mortality survey identified 144 deaths that occurred between 22 March 2020 and 11 July 2020. Among these, 47 (33%) matched the U.S. CDC definition for suspected COVID-19. Eleven (8%) deaths were attributed to other causes (including measles, tetanus, injury, drowning, cardiac arrest, childbirth, and foodborne illness or poisoning), and the remaining 86 (60%) deaths were of indeterminate causes. It is possible some of the indeterminate deaths were COVID-19-related, but data were insufficient to define a cause. We estimate the deaths captured by the survey represent a mean weighted estimate of 364 suspected COVID-19 and 523 indeterminate deaths across the full Rohingya population in the Kutupalong-Balukhali camps (see *Supplemental Materials*). Validation of these suspected COVID-19 deaths indicated consistency with the expected age distribution of COVID-19 deaths, and timing corresponded well with other events and data. No major issues were identified (see *Supplementary Materials 2*.*2*).

### Inferring infections from expected IFR and deaths

Based on the age distribution of the 34 camps and published age-specific IFR estimates, we estimate an expected median overall IFR of 0.096% (95% CI, 0.084-0.112%), with slight variability between camps (median range: 0.082% to 0.107%). Assuming these IFR estimates, we estimate the six reported confirmed COVID-19 deaths represent 6,220 SARS-CoV-2 infections (Table 1). Including the 47 survey deaths meeting the CDC case definition in addition to the 6 confirmed deaths (excluding one overlapping death), the number of infections increases to 53,907, though this represents only a random sample of households. Using the population-weighted estimate of 364 deaths plus the six confirmed deaths, we estimate 383,568 infections may have occurred by July, with nearly half (44.3%) of the population having been infected by that time. With six confirmed deaths out of the total 370 estimated, and 57 confirmed cases, this would indicate that 1 in 6,729 infections and 1 in 62 deaths were reported.

**Table 1.** Deaths by criteria and associated estimated infections, March – July 2020, Rohingya refugees, Kutupalong-Balukhali.

| Death criteria | N* | Estimated IFR,<br>per 1000<br>median (95% CI) | Estimated Infections<br>median (95% CI) | Percent Infected<br>median (95% CI) |
| --- | --- | --- | --- | --- |
| Confirmed | 6 | 0.96<br>(.84-1.12) | 6,220 (5,370-7,135) | 0.7% (0.6-0.8%) |
| Confirmed + Suspected** | 52 |  | 53,907 (46,540-61,841) | 6.2% (5.4-7.1%) |
| Confirmed + Weighted Suspected** | 370 |  | 383,568 (331,151-440,021) | 44.3% (38.2-50.8%) |

\*Deaths occurring during March - July 2020
\*\*Excludes 1 overlapping death

### Inferring cases and deaths from available data

The three scenarios, each using the same set of reported data on the weekly number of SARS-CoV-2 PCR tests and positives, and the resulting positivity rates, resulted in widely different epidemic shapes and sizes.

#### Scenario 1. Care seeking proportional to infection (Null model)

In Scenario 1, infections follow a similar epidemic curve shape to the reported cases from which they are inferred (Figure 4). Two waves are observed, but with only one major week of transmission occurring during the first wave (week 21). Like the reported cases, the majority of estimated infections are concentrated in the second wave.

**Figure 3.**
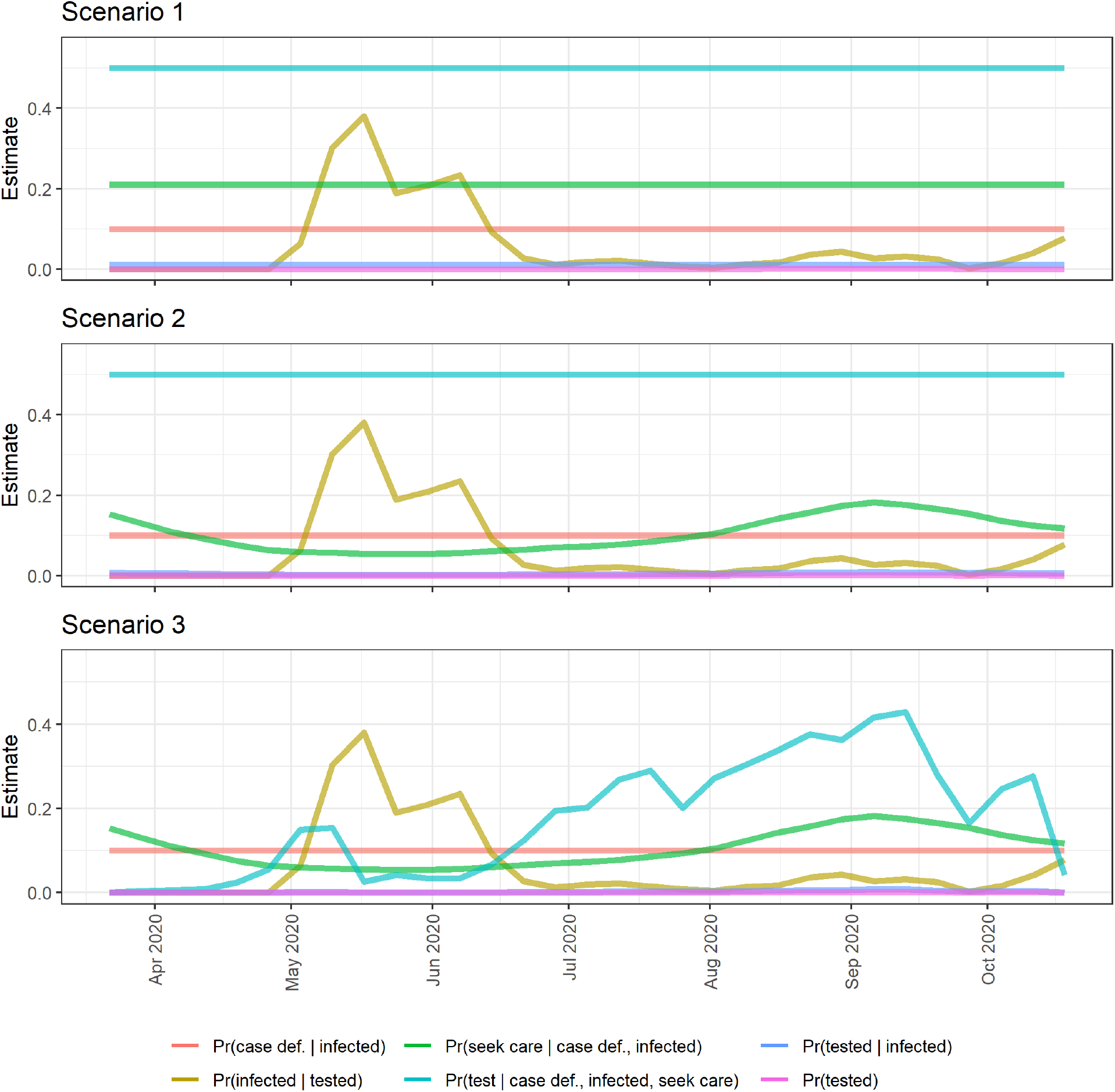
Probabilities serving as inputs into and estimated from the inference model, Scenarios 1-3. Each scenario assumes a set of probabilities derived from the scenario assumptions. All scenarios assume the same probability over time that a person in the population is tested, Pr(tested), calculated as the number of tests performed divided by total population (pink), the same probability of meeting the case definition given infected, Pr(case def. | infected) = 0.10 (orange), and the same probability of being infected among those tested (yellow), calculated as the test-positivity rate adjusted for test sensitivity. For Scenario 1, reported cases are assumed proportional to true infections over time, thus both probabilities of being tested (blue) and seeking care (green) assuming an individual is infected and meets the case definition are defined to be constant over time. For Scenario 2, we use a time varying estimate of the probability of seeking care (Pr(seek care | case def., infected), turquoise) derived from EWARS data. For Scenario 3, we also use a time-varying probability of being tested among infected individuals who seek care and meet the case definition (turquoise), Pr(test | case definition, infected, seek care).

**Figure 4.**
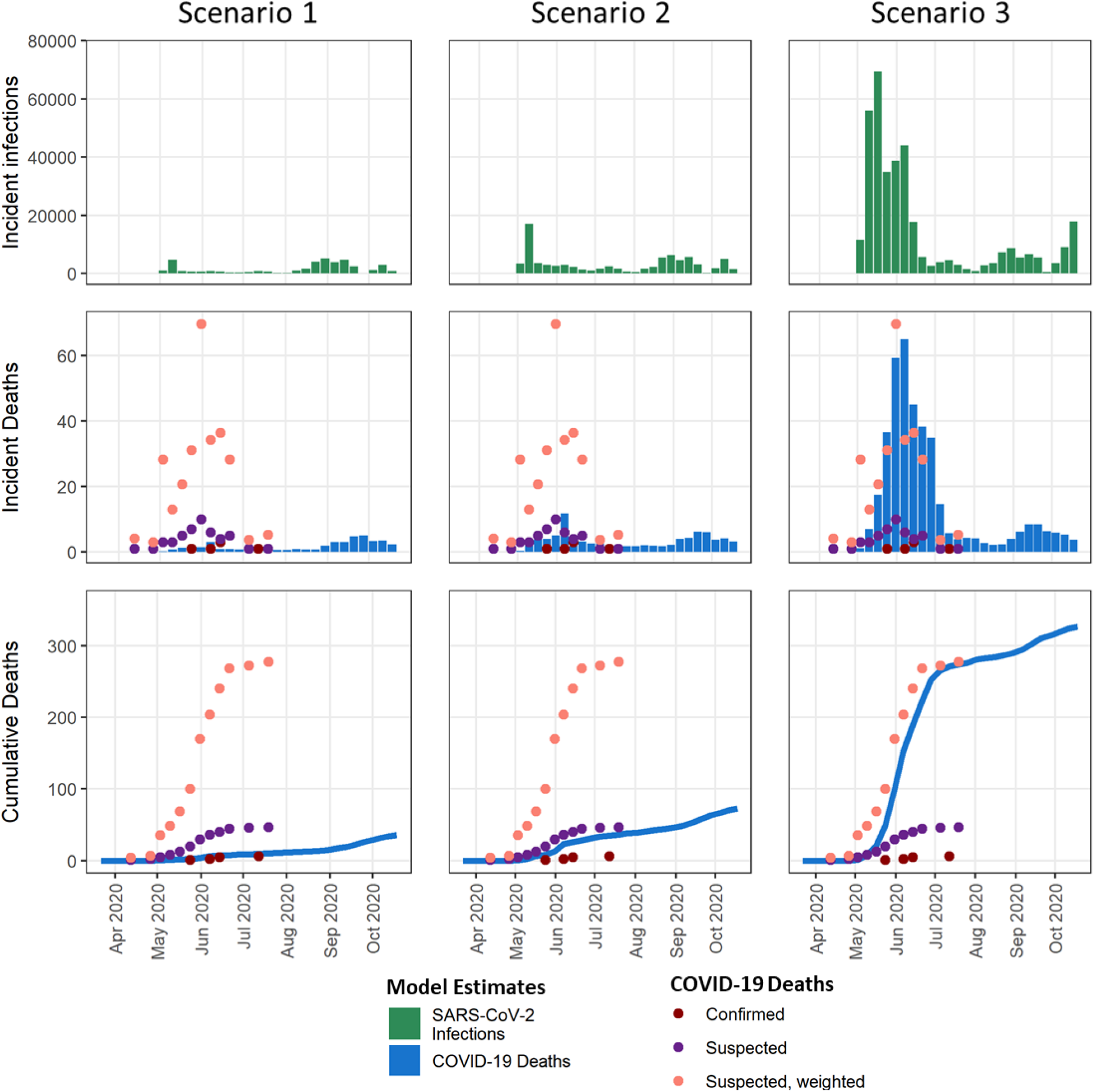
Inferred estimates of SARS-CoV-2 infections and deaths from testing data, Rohingya refugees, March-October 2020. Infections were estimated from the weekly counts of reported PCR-positive COVID-19 cases and number of RT-PCR tests performed, and test-positivity rate, along with assumed probability estimates (*see Figure 3 and Table S1)*. For Scenario 1, we assume case-seeking behavior and testing probability are static. Scenario 1 represents a situation where we believe the reported counts of cases and deaths are proportional to the level of infections in the population, similar to how we use cases and deaths in other populations around the globe. As such, under this scenario, the larger number of PCR-positive cases identified during weeks 35-40 represents a larger second wave with about 25,000 infections. For Scenario 2, testing probability remains constant, but care-seeking is assumed to be dynamic, and is defined by the ratio of observed ARI cases to expected ARI in this population over time using EWARS data. Under this second scenario, confirmed cases and PCR testing data produce a slightly larger overall disease burden than Scenario 1, with more infections concentrated in the first wave and a similar second wave. Scenario 3 allows both care seeking and testing to be dynamic, with testing probability estimated using the ratio of the number of negative tests to the baseline level of ARI in this population. Under this scenario, the official data represents an explosive initial outbreak and continued transmission during the later period. Applying the age-adjusted infection fatality ratio estimated for the Rohingya population using age-specific probability of death from the O’Driscoll et al., we estimate the number of deaths associated with these inferred infections^25^. We find similar timing, shape, and scale in the inferred deaths from Scenario 3 as compared to the sampling weight-adjusted deaths estimated to have occurred during March-July 2020 from the mortality survey (pink).

Comparing estimated deaths to those reported, this scenario resembles the confirmed deaths occurring during the first wave; though we would expect to see an additional 18 deaths occurring during a second wave of this magnitude, only 4 were reported. Under the assumptions of this scenario, the reported data represent an estimated 36,169 cumulative infections and 34 cumulative deaths occurring from May-October 2020.

#### Scenario 2. Time-varying care seeking

We observe that ARI cases, and the resulting *α*_*t*_, began decreasing in the week of 5 April 2020, shortly after cases began increasing (Figures S1, 3B). In Scenario 2, assuming these sudden changes in ARI approximate changes in care seeking over time, the official case numbers and testing data represent a notably different picture of the outbreak compared to Scenario 1. These data describe a larger first wave of infection, followed by low levels of continued transmission or introduction of SARS-CoV-2, and a similar fall wave of infection to Scenario 1 (Figure 4). Inferred deaths are substantially greater than the confirmed reported COVID-19 deaths throughout the time period, yet do not capture the magnitude implied by the mortality survey. Under the assumptions of this scenario, the reported data represent an estimated 72,363 cumulative infections and 67 cumulative deaths occurring from May-October 2020.

#### Scenario 3. Time-varying care seeking and testing

In Scenario 3, we derived the probability that an infected individual who seeks care is tested directly from ARI data. This probability increases over time from <5% early in the pandemic to 41% by mid-September (Figure 3C). Under these assumptions, reported case numbers and testing data estimate a large outbreak during the first wave, with the vast majority of infections occurring during May-July 2020 (80%), and only low levels of transmission for the remainder of the study period (Figure 4). Estimated deaths correspond well with the mortality survey-implied number of COVID-19 deaths. Under the assumptions of this scenario, we estimate the reported data may represent as many as 406,000 cumulative infections and 371 cumulative deaths occurring from May-October 2020 (Figure 4).

## Discussion

### Underrealized COVID-19 impacts

Critical gaps remain in our understanding of how much SARS-CoV-2 transmission has occurred and what the true COVID-19 impacts on morbidity and mortality have been in populations around the world. This is even more prominent among many of the world’s most vulnerable populations, including emergency-affected populations such as refugees and internally displaced persons. Many have pointed to limited testing capacities and poor contact tracing and surveillance systems as major drivers of this lack of documented impact ^28,29^. While these reasons may contribute to some knowledge gaps, they do not fully explain why health care systems have not been overwhelmed as was expected early in the pandemic ^30^. For vulnerable populations like the Rohingya refugees, where testing capacity has been mostly sufficient but willingness to seek care and be tested has been low, significant challenges exist to characterize and understand the virus’ transmission and impact. Better characterization of infection and disease transmission in these populations is essential to improve our response to the current pandemic and prepare for future epidemics and pandemics among vastly different contexts.

One reason for the underrealized COVID-19 impacts for populations living in low-income countries is likely that their probabilities of severe disease and death per infection were initially overestimated. This overestimation resulted from reliance on early estimates of age-specific mortality from small populations with incomplete information on infection and risk factors, prior to the availability of serology. Indeed, our revised IFR estimate of 0.096% is about one-third of what we previously estimated (0.36%) for the Rohingya population ^30^. Other factors, such as lower prevalence of comorbidities compared to other countries and potential cross-protection from previously circulating coronaviruses, may also have contributed to the reduced COVID-19 impact, but evidence on these is still lacking.^30–33^ Continued scientific investigation amongst populations in varied contexts is necessary for understanding the differential effects of SARS-CoV-2 infection and to support evidence-based decision-making and resource prioritization for insecure and vulnerable populations.

Even with lower severity and mortality rates, we would expect to have observed significantly more deaths than have been reported to date among the Rohingya. Numerous examples have demonstrated the possibility of unidentified large-scale transmission, including in Dharavi and Dhaka, where populations believed to have been minimally impacted by the pandemic were found to have substantial SARS-CoV-2 seroprevalence^11–13^. Indeed, it is likely that something similar occurred among the Rohingya. Although only serological testing will provide a direct measurement of the proportion infected, given current data and our model and examination of deaths through the mortality survey, these results provide have gained insight into how much transmission potentially occurred, mirroring what has been noted in other similarly population dense, low-resource settings.

Throughout this pandemic, public health experts and authorities have relied heavily on testing data to understand the course of the pandemic. While high rates of asymptomatic or mild infection coupled with insufficient capacity to test lead us to assume reported cases only represent a fraction of true infections in a population, the Rohingya population presents an additional challenge – active avoidance of testing and health care seeking, and variation in this behavior over time. Documented in multiple accounts, there were active, community-wide efforts to conceal both past and current symptoms that could indicate COVID-19 disease, which may be reflected by the dramatic decrease in care-seeking during the initial period of the pandemic ^18^. This likely stemmed from rumors and fear of being put into isolation, being forcibly moved to an island, or even being killed, coupled with a long history of violent persecution ^34–36^. In addition to multiple reported and personal communications indicating people were not willing to seek care, we also noted a dramatic sudden drop in all ARI cases reported. It is possible, however, that some of this decline was a result of the movement restrictions and lockdown measures actually reducing transmission with lockdown measures in place in Bangladesh during 23 March 2020 to 30 May 2020. In addition to the evidence that care-seeking declined dramatically, there is evidence of under-testing, particularly early in the pandemic. Estimates of the increasing probability of testing in *Scenario 3* mirror others reported for this population, which have suggested the willingness to be tested among those seeking care could have been as low as 5-10% early in the pandemic, and only reaching 51% after efforts to increase testing acceptance were implemented though testing capacity was available^20^. As policies changed over time, including a shift from required isolation of confirmed cases in isolation units to allowing at-home isolation, and as humanitarian organizations worked to establish trust and dispel rumors, care seeking and willingness to be tested appears to have changed over time.

### Implications of transmission using multiple data sources

We explored use of multiple distinct datasets that relate to the SARS-CoV-2 pandemic and COVID-19 disease in the Rohingya refugee population. By triangulating these data through inference modeling, we estimate under *Scenario* 3 that nearly one-half of the total Rohingya population, approximately 325,000 persons, may have been infected during a large epidemic peak from May-July 2020, and as many as 400,000 may have been infected from May-October 2020. We find these estimates approximate separate estimates based on extrapolation from survey-identified suspected COVID-19 deaths. While both approaches and data sources have limitations, their results independently pointed to similar timing and magnitude of infections and deaths in this population; both approaches were in support of *Scenario* 3, that a large outbreak occurred early in the pandemic and was largely unidentified at the time due to limited and variable care seeking and test seeking.

The suspected outbreak appears to coincide with Ramadan, which lasted from 25 April to 24 May 2020 (Figure 2B). Survey deaths started increasing three weeks after the start of Ramadan, which would place the time of infection for those deaths at the beginning of Ramadan. We know from anecdotal reports that the camps had largely been in lockdown up to Ramadan, but after pressure, mosques were allowed to reopen for the month. Additionally, during Ramadan, it is common practice for people to congregate in large groups to consume their evening meal and some travel to different mosques, all potentially contributing to increased population mixing, contact, and transmission during that time period. This estimated epidemic curve also corresponds well with that of the host community, which experienced a major outbreak during May-July 2020 ^9^.

Each of the data sources used in this analysis has strengths and limitations. The mortality survey provides a valuable resource for understanding potential unidentified COVID-19 deaths, from which transmission may be inferred. In a vulnerable population like this, where active efforts to hide illness and avoid care and testing were likely to have occurred, culturally appropriate approaches, as were used in this survey, may better elicit the truth than more traditional surveillance methodologies; understanding what was locally relevant and leveraging the community trust of the *mazis* was critical for attaining ground truth data in the absence of healthcare-seeking. Additionally, through specific data on the symptoms and reported causes, as well as the ages of these deaths, we can assess both classification of COVID-19 through case definitions and how they correspond with characteristics predictive of mortality from COVID-19, namely older age. These data also present several important limitations, specifically the uncertainty and potential for bias resulting from the lack of confirmatory testing and from secondary reporting. Without testing data and only limited symptom and disease data, misclassification of suspected COVID-19 deaths is possible. In the mortality survey, 21 deaths were reported as having “asthma” or “tuberculosis” as symptoms, and it is possible these were indications for difficulty breathing or shortness of breath, which would indicate suspected COVID-19. In fact, a portion of the 60% of deaths that remained unclassified may be attributable to COVID-19. This challenge is not isolated to the mortality survey - among 31 EWARS reports evaluated from 9 March to 11 October 2020, 68.5% (381/556) of deaths reported across the Rohingya camps were classified as “Other”. Though better cause of death characterization, during and outside of pandemic settings, is the focus of several long-term research efforts, this pandemic has magnified this challenge, with reports of people purposefully misrepresenting illnesses and causes of death^37^.

Improved sources of data to better assess care-seeking behavior and testing willingness are critical to using methods exemplified in this paper. Here we relied on proxy data through ARI reporting and published estimates, however, more accurate estimation of infection might be possible through well implemented quantitative and qualitative studies. As a result of our reliance on limited data, we cannot express statistical confidence in our quantitative estimates of cases and deaths that were inferred from the three scenarios. Nonetheless, we are confident that the reported data can tell a substantially different story when one accounts for care-seeking and testing behaviors. While we cannot definitively estimate the amount of transmission that has occurred or the number of true COVID-19-related deaths, the current narrative that the Rohingya have somehow been minimally infected with SARS-CoV-2 is likely underestimating the true burden of COVID-19. Instead, these data indicate a large, early, and unidentified outbreak occurred among the Rohingya refugees. As this pandemic continues to evolve, with novel, more transmissible variants and potential immune waning, understanding previous transmission is critical for understanding the risk remaining in populations like the Rohingya.

## Data Availability

Data necessary to conduct the primary analyses have been aggregated for sharing and are available with all code required on the github repository.

https://github.com/HopkinsIDD/rohingyaCOVID19burden

## Funding

This study was supported by a grant from the U.S. Centers for Disease Control and Prevention (grant number CDC-RFA-GH16-171902CONT19).

## Declaration of Competing Interest

The authors declare that they have no known competing financial interests or personal relationships that could have appeared to influence the work reported in this paper.

## Acknowledgments

We would like to acknowledge the International Organization for Migration, Daniel Coyle (IOM), and the Rohingya researchers who conducted the mortality survey and shared those data; the United Nations High Commissioner for Refugees for providing data on camp population as well as insights into the population and situation on the ground; Médecins Sans Frontières for contributing insight and data; members of the Johns Hopkins Infections Disease Dynamics group and Johns Hopkins Center for Humanitarian Health for insight and feedback during the development of this project; and the U.S. Centers for Disease Control and Prevention for funding this work and providing valuable input and insight.

## Notes

### Competing Interest Statement

The authors have declared no competing interest.

### Author Declarations

This work was conducted as part of public health response and was conducted under IRB exemption from Johns Hopkins Bloomberg School of Public Health as "public health surveillance" activities, IRB00011834. Approval for use of the mortality survey data was granted by the International Organization for Migration. Approval for use of refugee camp population data for the Rohingya refugees was provided by the United Nations High Commissioner for Refugees. Approval for use of MSF facility data was provide by Medecins Sans Frontieres. All other data used in this analysis are publicly available.

## References

1 Truelove S, Abrahim O, Altare C, et al. The potential impact of COVID-19 in refugee camps in Bangladesh and beyond: A modeling study. PLOS Med 2020; 17: e1003144.

2 Anwar S, Nasrullah M, Hosen MJ. COVID-19 and Bangladesh: Challenges and How to Address Them. Front Public Health 2020; 8. DOI:10.3389/fpubh.2020.00154.

3 Raju E, Ayeb-Karlsson S. COVID-19: How do you self-isolate in a refugee camp? Int J Public Health 2020; 65: 515–7.

4 Vonen HD, Olsen ML, Eriksen SS, Jervelund SS, Eikemo TA. Refugee camps and COVID-19: Can we prevent a humanitarian crisis? Scand J Public Health 2021; 49: 27–8.

5 COVID-19 Isolation and Treatment Centre opens for Bangladeshi communities and Rohingya refugees in Cox’s Bazar. ReliefWeb. 2020; published online Sept 7. https://reliefweb.int/report/bangladesh/covid-19-isolation-and-treatment-centre-opens-bangladeshi-communities-and-rohingya (accessed March 1, 2021).

6 Rodion Ebbighausen. In Rohingya refugee camps, coronavirus is under control — for now. DW.com. 2020; published online June 19. https://www.dw.com/en/in-rohingya-refugee-camps-coronavirus-is-under-control-for-now/a-53873931 (accessed March 1, 2021).

7 Shehab Sumon. Rohingya refugee camps braced for coronavirus. Arab News. 2020; published online March 2. https://arab.news/zmpxf (accessed March 1, 2021).

8 Agence France-Presse. First coronavirus case at Rohingya refugee camps in Bangladesh. The Guardian. 2020; published online May 14. http://www.theguardian.com/world/2020/may/14/first-coronavirus-case-rohingya-refugee-camps-bangladesh (accessed Dec 6, 2020).

9 COVID-19 Dashboard. WHO Coxs Bazar Data Hub. https://cxb-epi.netlify.app/post/covid-19-dashboard/ (accessed Nov 20, 2020).

10 Kucharski AJ, Klepac P, Conlan AJK, et al. Effectiveness of isolation, testing, contact tracing, and physical distancing on reducing transmission of SARS-CoV-2 in different settings: a mathematical modelling study. Lancet Infect Dis 2020; 20: 1151–60.

11 Soutik Biswas. How Asia’s biggest slum contained the coronavirus. BBC News. 2020; published online June 22. https://www.bbc.com/news/world-asia-india-53133843 (accessed Dec 6, 2020).

12 India coronavirus: ‘More than half of Mumbai slum-dwellers had Covid-19’. BBC News. 2020; published online July 29. https://www.bbc.com/news/world-asia-india-53576653 (accessed Dec 6, 2020).

13 Tawsia Tajmim. Dhaka slum dwellers develop herd immunity. Bus. Stand. 2020; published online Oct 12. https://tbsnews.net/coronavirus-chronicle/covid-19-bangladesh/45-dhaka-population-develops-covid-19-antibodies-144205 (accessed Dec 6, 2020).

14 Malani A, Shah D, Kang G, et al. Seroprevalence of SARS-CoV-2 in slums versus non-slums in Mumbai, India. Lancet Glob Health 2021; 9: e110–1.

15 George CE, Inbaraj LR, Chandrasingh S, Witte LP de. High seroprevalence of COVID-19 infection in a large slum in South India; what does it tell us about managing a pandemic and beyond? Epidemiol Infect 2021; 149. DOI:10.1017/S0950268821000273.

16 Figar S, Pagotto V, Luna L, et al. Community-level SARS-CoV-2 Seroprevalence Survey in urban slum dwellers of Buenos Aires City, Argentina: a participatory research. medRxiv 2020; : 2020.07.14.20153858.

17 McCulloh I, Kiernan K, Kent T. Inferring True COVID19 Infection Rates From Deaths. Front Big Data 2020; 3. DOI:10.3389/fdata.2020.565589.

18 The Stories being told: Rohingya report on the epidemic. International Organization for Migration, 2020 https://www.acaps.org/special-report/bangladesh-covid-19-explained-rohingya-report-epidemic-stories-being-told (accessed Nov 20, 2020).

19 Mwananyanda L, Gill CJ, MacLeod W, et al. Covid-19 deaths in Africa: prospective systematic postmortem surveillance study. BMJ 2021; 372: 334.

20 McGowan CR, Hellman N, Chowdhury S, Mannan A, Newell K, Cummings R. COVID-19 testing acceptability and uptake amongst the Rohingya and host community in Camp 21, Teknaf, Bangladesh. Confl Health 2020; 14: 74.

21 Lopez-Pena P, Davis CA, Mobarak AM, Raihan S. Prevalence of COVID-19 symptoms, risk factors, and health behaviors in host and refugee communities in Cox’s Bazar: a representative panel study. nCoV, 2020 DOI:10.2471/BLT.20.265173.

22 Rohingya emergency. UNHCR. https://www.unhcr.org/rohingya-emergency.html (accessed Nov 20, 2020).

23 Rohingya Refugees Population by Location at Camp and Union Level - Cox’s Bazar Humanitarian Data Exchange. Humanit. Data Exch. https://data.humdata.org/dataset/site-location-of-rohingya-refugees-in-cox-s-bazar (accessed March 22, 2021).

24 Early Warning, Alert and Response System (EWARS). World Health Organization https://www.who.int/bangladesh/emergencies/Rohingyacrisis/ewars (accessed Dec 4, 2020).

25 O’Driscoll M, Ribeiro Dos Santos G, Wang L, et al. Age-specific mortality and immunity patterns of SARS-CoV-2. Nature 2021; 590: 140–5.

26 The IEDCR and partners share insights on the prevalence, seroprevalence and genomic epidemiology of COVID-19 in Dhaka city. Icddrb - Press Releases. 2020; published online Oct 12. https://www.icddrb.org/quick-links/press-releases?id=97&task=view (accessed March 15, 2021).

27 COVID-19 Status for Bangladesh. https://idare.maps.arcgis.com/apps/MapSeries/index.html?appid=e621575c559e4b5185b56382071ed184 (accessed May 17, 2021).

28 Dahab M, van Zandvoort K, Flasche S, et al. COVID-19 control in low-income settings and displaced populations: what can realistically be done? Confl Health 2020; 14: 54.

29 Poole DN, Escudero DJ, Gostin LO, Leblang D, Talbot EA. Responding to the COVID-19 pandemic in complex humanitarian crises. Int J Equity Health 2020; 19: 41.

30 Truelove S, Abrahim O, Altare C, et al. The potential impact of COVID-19 in refugee camps in Bangladesh and beyond: A modeling study. PLOS Med 2020; 17: e1003144.

31 Yaqinuddin A. Cross-immunity between respiratory coronaviruses may limit COVID-19 fatalities. Med Hypotheses 2020; 144: 110049.

32 Ma Z, Li P, Ji Y, Ikram A, Pan Q. Cross-reactivity towards SARS-CoV-2: the potential role of low-pathogenic human coronaviruses. Lancet Microbe 2020; 1: e151–e151.

33 Pinotti F, Wikramaratna PS, Obolski U, et al. Potential impact of individual exposure histories to endemic human coronaviruses on age-dependent severity of COVID-19. BMC Med 2021; 19: 19.

34 Health behaviours & COVID-19. ACAPS, 2020 https://www.humanitarianlibrary.org/resource/rohingya-response-health-behaviours-covid-19 (accessed March 1, 2021).

35 Nazmun Naher Shishir. In Bangladesh, internet restrictions, rumours worsen COVID-19 fears in Rohingya camps. Firstpost. 2020; published online June 23. https://www.firstpost.com/long-reads/in-bangladesh-internet-restrictions-rumours-worsen-covid-19-fears-in-rohingya-camps-8514181.html (accessed March 1, 2021).

36 Poppy McPherson RP. Fear stops Rohingya getting tested as virus hits refugee camps. Reuters. 2020; published online June 5. https://www.reuters.com/article/us-health-coronavirus-rohingya-refugees-idUSKBN23C1GV (accessed March 1, 2021).

37 Child Health and Mortality Prevention Surveillance (CHAMPS). champshealth.org. https://champshealth.org/ (accessed March 4, 2021).

