## Supplemental Materials for "Estimating the Burden of SARS-CoV-2 among the Rohingya Refugees"

### 1. METHODS

#### 1.1. Infection fatality ratio

The infection fatality ratio (IFR) was estimated by as population-weighted mean of age-specific probability of death among infected individuals. Estimates for probability of death among infected individuals, $\Pr(death|infection)$ were derived from work done by O'Driscoll et al. which used serosurvey data from across 45 countries to estimate the age-specific probabilities of death.[1] We used the estimated ensemble median and 95% confidence interval from that work and applied to the sub-block-specific age-specific populations as follows:

$$IFR_{subblock} = \sum_{a} \Pr(death|infection)_a * \frac{population_a}{population_{total}}$$

Where $a$ represents age group. To get the camp-level and overall IFR estimates, the population-weighted means were estimated in a similar way, accounting for the population proportion of the sub-blocks and camps, respectively. Population data were acquired from the United Nations High Commissioner for Refugees (UNHCR).

Given the age distribution of each of the 29 camps, we estimate the overall infection fatality ratio (IFR) should fall within the range of 0.029% to 0.183%, with a median expected IFR of 0.096% (95% CI, 0.084-0.112%). At the camp level, notable variation was found, ranging from a median IFR estimate of 0.082% in Kutupalong RC to 0.107% in Camp 15. Based on these estimates, with 100% attack rate among 860,697 Rohingya refugees, we would expect a median total of 830 (95% CI, 724-962) deaths, with as few as 252 and as high as 1572 deaths.

#### 1.2. Infection and testing inference model

As described in the Methods of the main text, we use Bayes' Theorem to express the probability of infection in the population at a specific time-point as:

$$\Pr(\text{infected}) = \frac{\Pr(\text{infected} \mid \text{tested}) \Pr(\text{tested})}{\Pr(\text{tested} \mid \text{infected})} \tag{1}$$

<u>$Pr(infected)$</u>

The $Pr$(infected) is the probability that any individual in the population is infected during the specified time period. Once we estimate this value, we can apply it to the total population to estimate the number of infections at time $t$:

$$infections_t = \Pr(infected)_t \times population$$

$Pr$(infected | tested)

$Pr$(infected | tested) is the time-varying positivity rate, adjusting for test sensitivity (i.e., the probability that an individual tested is positive for SARS-CoV-2 infection).

$Pr(tested)$

$Pr$(tested) is calculated as the number of tests performed divided by the total population at each time-point (e.g., daily or weekly). We quantify this over time from the number of PCR-positive cases reported:

$$Pr(\text{infected})_t = n_{case}/population$$

Where $n_{case}$ is the time-series of PCR-confirmed, reported cases.

$Pr(tested \mid infected)$

We further assume that to get tested in this population, an individual must seek care and be willing to be tested ("seek care & testing"), present with symptoms that meet the case definition for COVID-19 ("case definition"), and there must be testing availability at the time of care seeking. For this population and analysis, we assume testing of asymptomatic individuals is negligible. We represent the probability that an infected individual is tested in the population as

$$\begin{aligned}\Pr(\text{tested} \mid \text{infected}) = \; & \Pr(\text{case definition} \mid \text{infected}) \times \\ & \Pr(\text{seek care \& testing} \mid \text{case definition, infected}) \times \\ & \Pr(\text{tested} \mid \text{case definition, infected, seek care \& testing})\end{aligned} \quad (2)$$

*$Pr$(case definition | infected)*

The first probability that we need to define to estimate the $\Pr(tested|infected)$ is the $Pr$(case definition | infected), or the probability that an infected individual meets the case definition for testing. This we can assume is a function of the probability of symptomatic disease and having symptoms that specified by a case definition for testing, such as fever and difficulty breathing or cough. Recent serosurveys from Dhaka, Bangladesh and Iceland have estimated only 10% and 50% of infected individuals experience COVID-19-related illness [2,3]. Assuming similarity between Dhaka and the Rohingya, for this analysis we use $Pr(\text{case definition} \mid \text{infected}) = 0.10$.[2]

*$Pr$(seek care & testing | case definition, infected)*

The probability that an individual sought care and testing, given they met the case definition and were infected. We again draw from the results of a serological survey in Dhaka, Bangladesh to provide an estimate for the probability of being detected given infection. This survey, conducted in between April and July 2020, found an estimated 45% seroprevalence in Dhaka, in a population of 8,900,000, equating to 4 million infections. However, during this time period only 38,324 confirmed cases were reported, giving an estimated $\Pr(\text{detected} \mid \text{infected})$=0.0096 [4]. Assuming an average sensitivity for RT-PCR of

0.80, we estimate $\Pr(\text{tested} \mid \text{infected})$=0.012 [5]. From this and *Equation 2* we can estimate an average probability that an infected individual who meets the case definition for test both seeks care and testing and is tested as

$$\Pr(seek\ care\ \&\ testing \mid\ case\ definition, infected) = \frac{\Pr(tested \mid infected)}{\Pr(case\ definition \mid infected) \times \Pr(tested \mid case\ definition, infected, seek\ care)}$$

In Bangladesh, a large portion of health care is provided outside of the standard healthcare providers (e.g., traditional healers, untrained, pharmacists); one study found 52% of parents sought care for children with diarrheal illness at standard care provider, with only 13% of seeking care from public health providers.[6] Assuming 60% of infected individuals seek care from standard providers, and thus receive testing for COVID-19, we estimate the proportion of symptomatic individuals seeking care and testing as:

$$\Pr(seek\ care\ \&\ testing \mid\ case\ definition, infected) = \frac{0.012}{0.10 \times 0.60} = 0.22$$

This value is similar to that from a study of street dwellers in Dhaka that found that only 22% of individuals seek care for acute respiratory infection (ARI) at a standard health facility.

*Time-variable care-seeking*

To explore the potential impact of changing care-seeking behavior, we used the acute respiratory illness (ARI) burden reported through EWARS reports from January 2018–February 2020 to define the expected ARI. We fit a linear regression model with both monthly and sequential weekly terms to these data, and from this projected the weekly expected ARI cases for March-December 2020. From these expected numbers we estimated the percent reduction in care seeking during 2020 (Figure S1). We assume that COVID-19 ARI cases should seek care with same proportion as non-COVID-19 ARI cases, assuming they are experiencing similar symptoms and disease, thus

$$Pr(\text{seek care \& testing} \mid \text{case definition, infected}) = Pr(\text{seek care \& testing} \mid \text{case definition}, \neg\text{infected})$$

For Scenario 2, we defined

$$Pr(seek\ care\ \&\ testing \mid \textit{case definition, infected})_t = Pr(seek\ care\ \&\ testing \mid \textit{case definition, infected})_0 * \alpha_t$$

Where $\alpha_t$ is the percent reduction estimated from EWARS data.

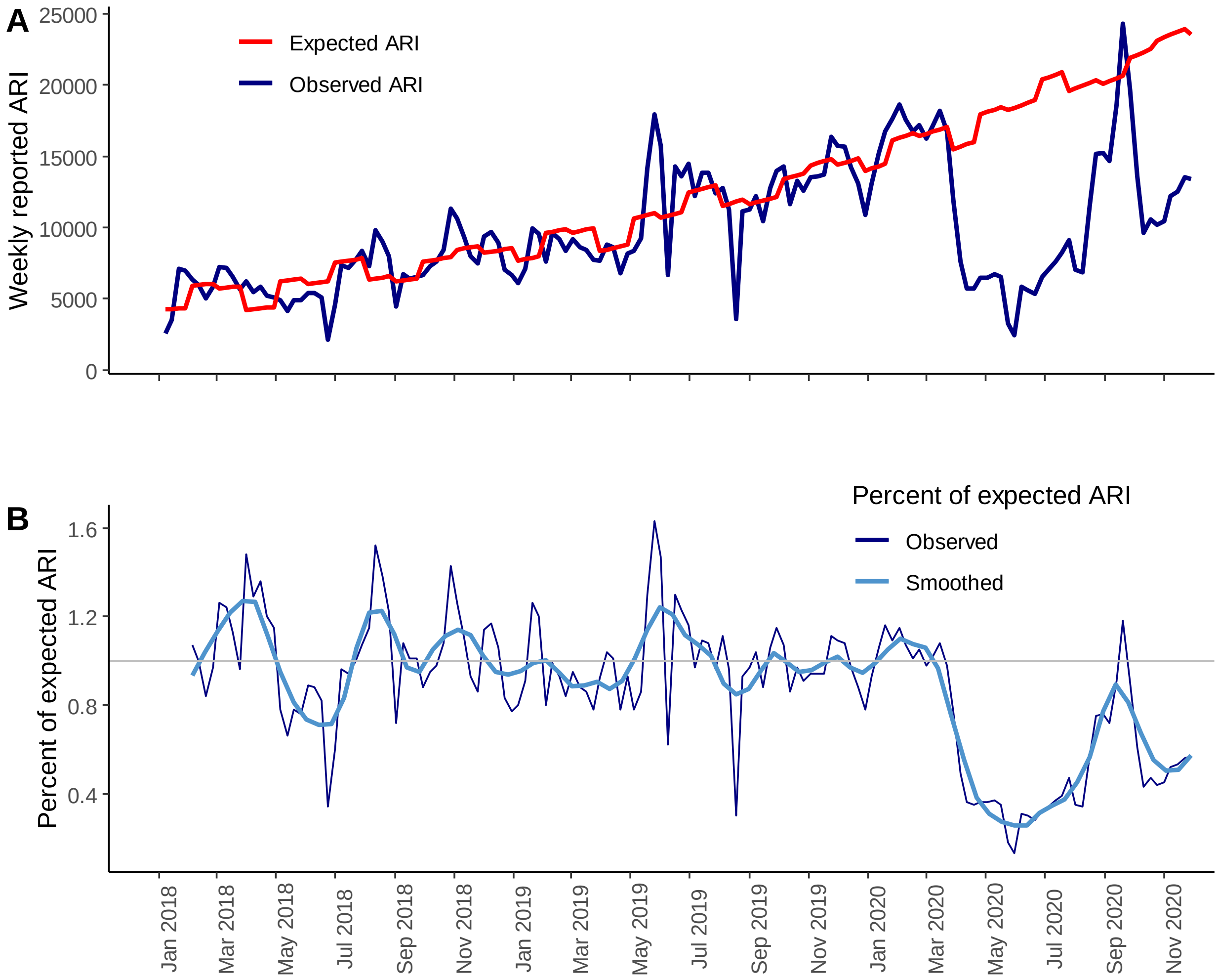


**Figure S1. Observed and expected acute respiratory infection and percent change from expected, Rohingya, 2018-2020.** (A) Data reported by community health workers through the Emergency Warning, Alert and Response System (EWARS) on acute respiratory infection (ARI) (navy blue) from January 2018 – February 2020 were used to fit a linear regression model accounting for week and month (red). (B) From these we estimated a percent change from the expected ARI. We found a sudden decline in ARI reporting starting the week of 5 April 2020 (Epiweek 15), dropping to 49% of the expected ARI numbers. At the lowest point, reported ARI was at 13% of the expected level, occurring the first week of June 2020 (Epiweek 23). Reporting appears to have steadily increased towards expected levels and as of the first week of December 2020 was at 57%.

<u>$Pr(tested \mid case\ definition, infected, seek\ care\ \&\ testing)$</u>

We similarly assume that individuals experiencing ARI symptoms compatible with the case definition are tested same probability regardless of their true infection status. Thus

$$Pr(\text{tested} \mid \text{case definition, infected, seek care \& testing}) \\ = Pr(\text{tested} \mid \text{case definition, seek care \& testing})$$

For the static scenarios (Scenarios 1 & 2), we assumed $Pr(\text{tested} \mid \text{case definition, infected, seek care \& testing}) = 0.50$. This value was derived from McGowan et al., which found 50% of Rohingya patients seeking care between 9 July and 21 October 2020 accepted testing for SARS-CoV-2.[7]

#### 1.3. Evidence for limited testing among ARI patients.

Data provided by Médecins Sans Frontières (MSF) on ARI cases in two MSF facilities provides insight into testing limitations among the Rohingya. These data, which include only individuals designated as acute respiratory illness, indicate SARS-CoV-2 testing among ARI patients did not begin until the week of 31 May 2020 (epi week 23, n=1 tested), the 3 weeks after the first case of SARS-CoV-2 infection was detected among the Rohingya refugees (Figure S2). Testing did not increase to >90% of ARI cases tested for another 4 weeks. During the period of testing ramp-up, test-positivity was also observed to increase sharply, indicative of a surging outbreak: during the week of 14 June 2020 (epi week 25), test-positivity reached 26% (and 29% of ARI cases were reportedly not tested for SARS-CoV-2). This spike in test-positivity at these two MSF facilities occurred 3 weeks after the overall test-positivity in the Rohingya camps spiked to over 30%, occurring during the week of 24 May 2020, during which only 1 test was performed among ARI cases seen at MSF facilities (1 of 1 positive). According to our inference model and results from Scenario 1, the first wave of this epidemic may have been nearly over by the time testing was sufficiently being performed (Figure 4). Because MSF facility testing may not be fully generalizable to all testing across the full population, and because these data also include non-refugee individuals, these data were not used to directly inform the inference model in this analysis. However, they do serve to demonstrate empirical evidence of testing limitations over time at two of the health facilities serving the Rohingya refugee population during the COVID-19 Pandemic.

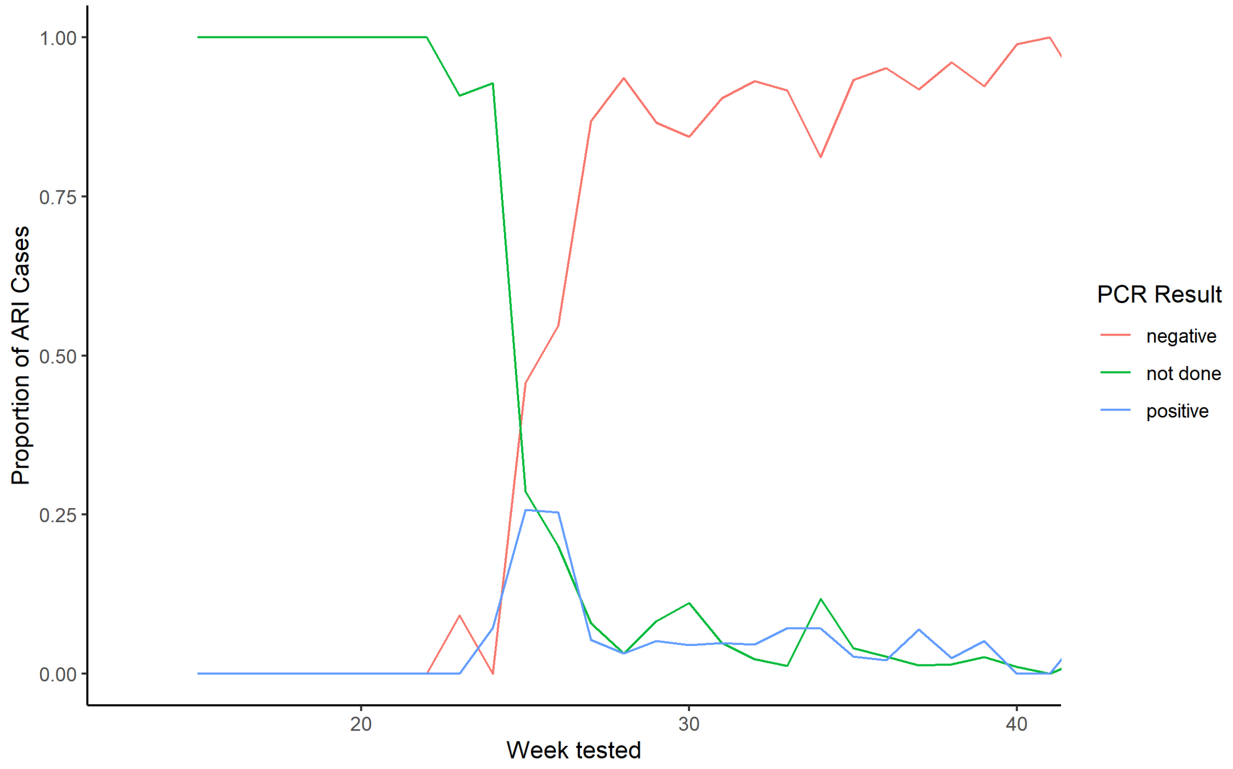


**Figure S2. Testing rate and test-positivity at two MSF health facilities in the Kutupalong-Balukhali refugee camps, April – October 2020.** Test status and result was reported for patients seeking care for acute respiratory illness (ARI). Proportions are presented for the week care and testing were sought.

#### 1.4. Scenario parameter input values

**Table S1. Scenarios and parameter assumptions used in the inference model.**

| Scenario | Time-varying care seeking | Time-varying testing | $\mathbf{Pr(seek\ care\ \&\ testing\ \vert\ case\ def., infected)}$* | $\mathbf{Pr(tested\ \vert\ seek\ care\ \&\ testing)}$ ** | $\mathbf{Pr(case\ definition\ \vert\ infected)}$ |
|---|---|---|---|---|---|
| ***Scenario 1*** | No | No | 0.22 | 0.60 | 0.10 |
| ***Scenario 2*** | Yes | No | 0.22* | 0.60 | 0.10 |
| ***Scenario 3*** | Yes | Yes | 0.22* | 0 - 0.41** | 0.10 |

**In the time-varying models, parameter estimates represent baseline values, prior to changes in care-seeking.*
***Varies over time, as quantified by EWARS ARI data.*

### 2. MORTALITY SURVEY DATA

The IOM mortality survey collected data on a total of 144 deaths that were reported to occur between 1 April 2020 and 11 July 2020.

#### 2.1. COVID-19 case definitions

For this analysis, the U.S. CDC's case definition was used as it more functionally aligned with the data collected from the IOM mortality survey. In the survey, 6 specific symptoms were collected: (1) fever, (2) cough/sore throat, (3) difficulty breathing, (4) diarrhea, (5) vomiting, (6) muscle or body aches. And additional question was asked about other symptoms, though this was largely recorded as "No" or left blank or (48%). Only 16 (11%) individuals had actual symptoms reported for this question, the rest were co-morbid conditions or other comments. We used the 2020 Interim Case Definition (Interim-20-ID-01) from the CDC to define a s*uspected COVID-19 case (https://wwwn.cdc.gov/nndss/conditions/coronavirus-disease-2019-covid-19/case-definition/2020/)*.

The clinical criteria are as follows:

*At least two of the following symptoms: fever (measured or subjective), chills, rigors, myalgia, headache, sore throat, new olfactory and taste disorder(s)*

*OR*

*At least one of the following symptoms: cough, shortness of breath, or difficulty breathing*

*OR*

*Severe respiratory illness with at least one of the following:*

- *Clinical or radiographic evidence of pneumonia, OR*
- *Acute respiratory distress syndrome (ARDS).*

*AND*

*No alternative more likely diagnosis*

The clinical case definition from the WHO was explored, but was decided to be less aligned with how the data were collected, specifically due to grouping cough and sore throat and not specifically asking about additional symptoms, including weakness/fatigue, coryza, headache, or altered mental status, all of which are specific to the WHO case definition (https://www.who.int/publications/i/item/WHO-2019-nCoV-Surveillance_Case_Definition-2020.2). The clinical criteria for this definition, updated 16 December 2020 is as follows:

- *Acute onset of fever AND cough; OR*
- *Acute onset of ANY THREE OR MORE of the following signs or symptoms: Fever, cough, general weakness/fatigue, headache, myalgia, sore throat, coryza, dyspnoea, anorexia/nausea/vomiting, diarrhoea, altered mental status.*

Following these case definitions, 47 (33%) and 34 (24%) had sufficient data and met the CDC and WHO case definitions, respectively. 11 (8%) of deaths could be attributed to other causes with some certainty (including measles, tetanus, injury, drowning, cardiac arrest, childbirth, foodborne illness or poison), and 87 and 99 were of indeterminate cause with the CDC and WHO case definitions, respectively. It is possible many of the indeterminate deaths were COVID-19 related but the data were incomplete or not reported correctly due to the survey being retrospective and informed by secondary informants.

### 2.2. Alignment between mortality survey deaths and confirmed COVID-19 cases and deaths

Evaluation of the characteristics of the deaths in the mortality survey data reveals no obvious outliers or characteristics that would cause us to doubt these data. We find the deaths are evenly distributed between males and females (female: 50%), with no obvious trends over time (Figure S3a). No obvious trends over time were observed between time and camps in which the deaths occurred (Figure S3b), or which region of the megacamp (Figure S3c), though the deaths were identified first in the eastern side of the Kutupalong-Balukhali megacamp, occurring six weeks prior to the other regions. This fits with introduction based on entry points and population density. Cause of death was also evenly distributed over time (Figure S3d).

As of 21 October 2020, 317 laboratory-confirmed cases were reported among the Rohingya in Cox's Bazar, with the first case detected on 14 May 2020. Among these cases, 9 were confirmed as COVID-19-associated deaths. These cases have occurred in what appears to have been two waves (Figure 2A), the first of which corresponds in timing with the deaths captured through the mortality survey (Figure 2B).

Date of death was available for six of the confirmed deaths. These deaths were distributed over weeks 22, 24, 25, and 29, coinciding well with the timing of the survey deaths (Figure 2B). The confirmed deaths were distributed across camps 3, 10, 11, 14, and Kutupalong RC, demonstrating no obvious discrepancies from the survey deaths, with the exception of Kutupalong RC, in which the survey was not conducted (Figure 2C).

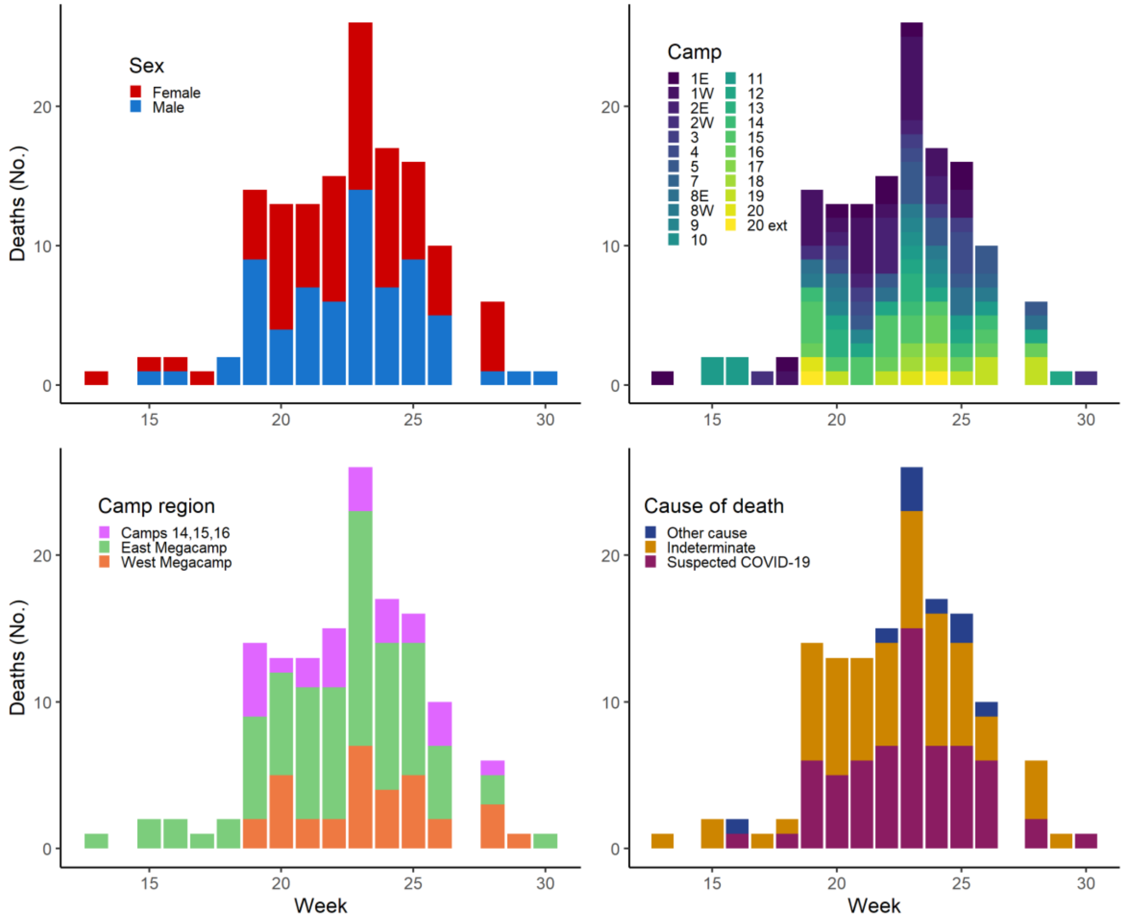


**Figure S3. Deaths by week, weeks 13-31, 2020.** These deaths include 144 deaths identified by the ACAPS mortality survey and 6 deaths reported with confirmed SARS-CoV-2. Sex distribution (A), camp of residence (B), camp region (C), and cause of death (D) indicate no major differences across time. There is some indication that deaths were occurring earliest in camps on the eastern side of the megacamp, potentially indicating the outbreak started there before it spread to the rest of the areas.

#### *2.2.1. Age distributions of cases and deaths*

As a result of the well-documented increasing severity by age among individuals infected with SARS-CoV-2, we expect deaths among the Rohingya to be similarly skewed toward older individuals, with those 65 and older comprising 66% of deaths from COVID-19, despite this age group on promising 2.5% of the total population (Figure S3). Comparing the age-specific expected number deaths (Figure 2B) to the age-specific distribution of the survey-identified deaths and the confirmed COVID-19 related deaths (Figure 2D), we see a notable similarity in the shapes of these distributions. Among the *suspected COVID-19* deaths, 41% (n=60) were among individuals aged 65 years and older. These distributions are in sharp contrast with the distribution of deaths by age identified in among the Rohingya in a 2018 survey, which more closely resembles that of the population overall[8]. In the 2018 survey, the individuals aged 61 years and older comprised 4% of the population surveyed and 17% of the deaths. One notable divergence between expected deaths and current deaths (Figures 2B and 2D) is among the 80+ age group. It is possible this is due to incorrect population denominators, reduced infection among older individuals, or some other survival factors. Similar reductions in mortality among the oldest age groups as compared to expected were observed in India[9].

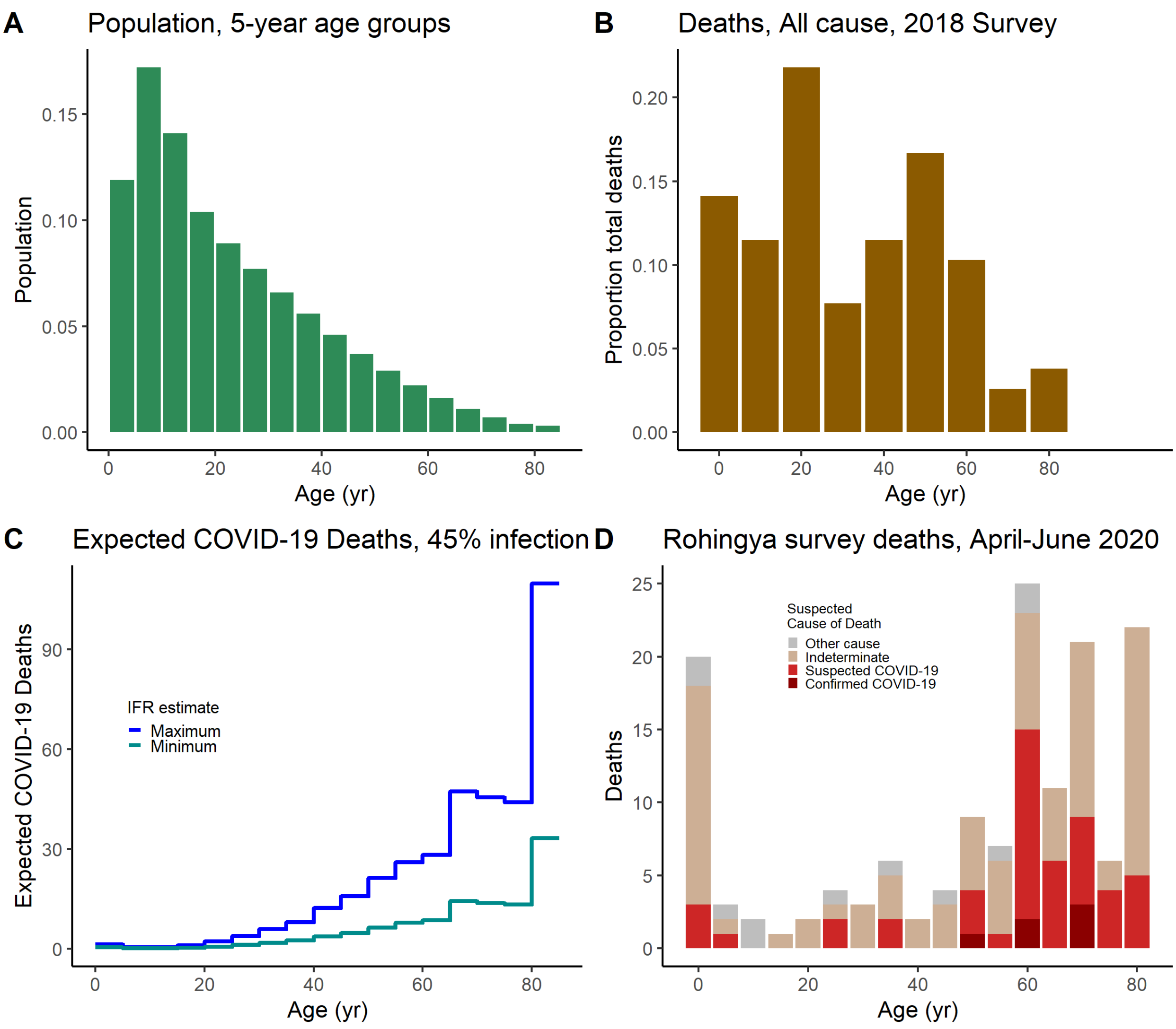


**Figure S3. Deaths, population, and age among the Rohingya refugees.** (A) Population age distribution, 5-year age groups. (B) Frequency of reported deaths among the Rohingya, during March 2017 to March 2018[8]. (C) The number of expected deaths from COVID-19 was estimated for each 5-year age group among the Rohingya, assuming 45% infection attack rate among a population of 860,697, which was found for Dhaka, Bangladesh from a serosurvey performed during April-June 2020. (D) IOM survey deaths and official confirmed COVID-19 deaths in the Rohingya during April-July 2020. IOM survey deaths are categorized by cause/suspicion as no testing was performed on these. The final age group, denoted as 80, represents 30 years, 80-110. The elevated deaths in the 80-110 age group is partly a result of it representing 30 years, in contrast with 5 years among the other age groups.

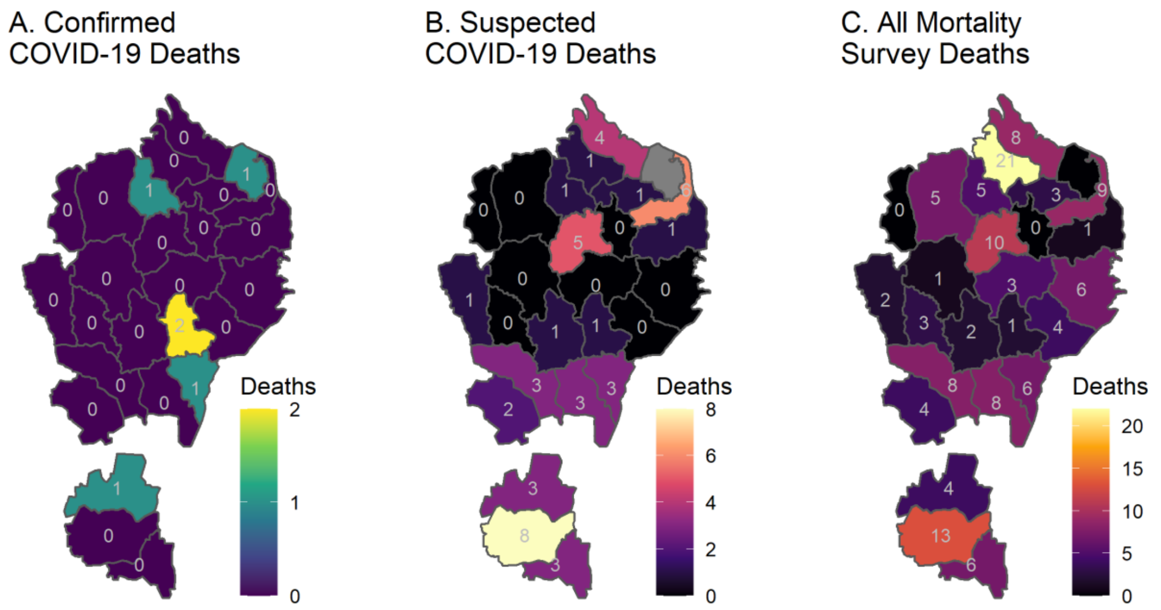


**Figure S4. Deaths by camp among the Rohingya, April-July 2020.** A. Reported laboratory-confirmed COVID-19-related deaths. B. Survey identified deaths classified as *suspected COVID-19* according to the CDC case definition. C. All unreported deaths identified through the mortality survey conducted by the International Organization for Migration and Rohingya researchers.

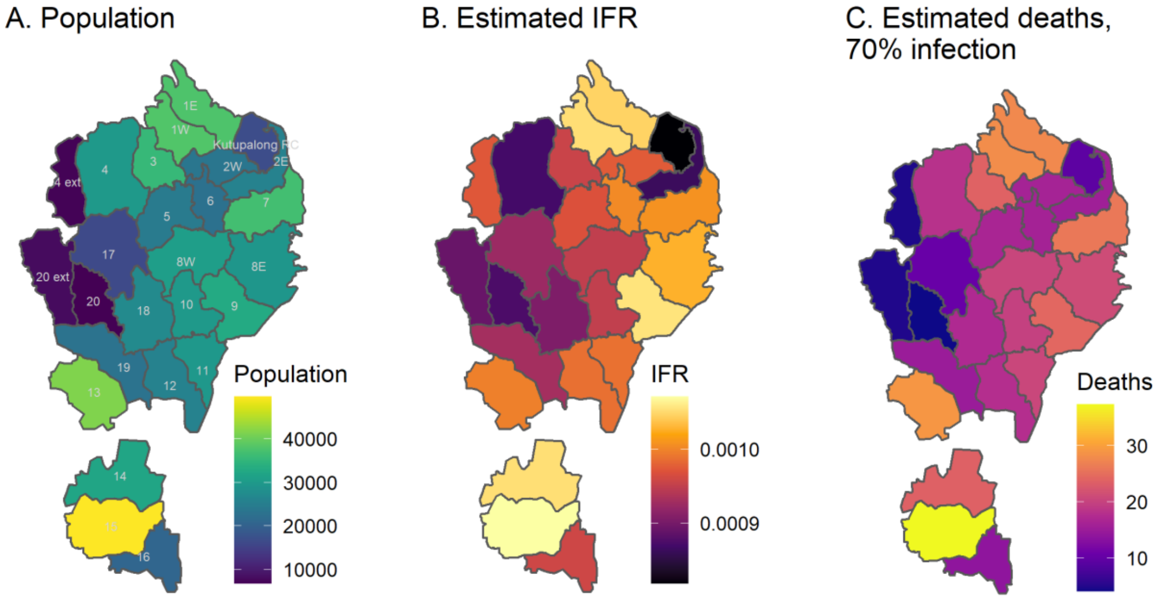


**Figure S5. Camp-specific population, infection fatality ratio, and estimated number of deaths assuming 70% cumulative infection.** Total population for each camp, median estimated the camp-specific IFR, and estimated cumulative deaths estimated are from the camp age distributions and estimates for the median age-specific fatality probability[1].

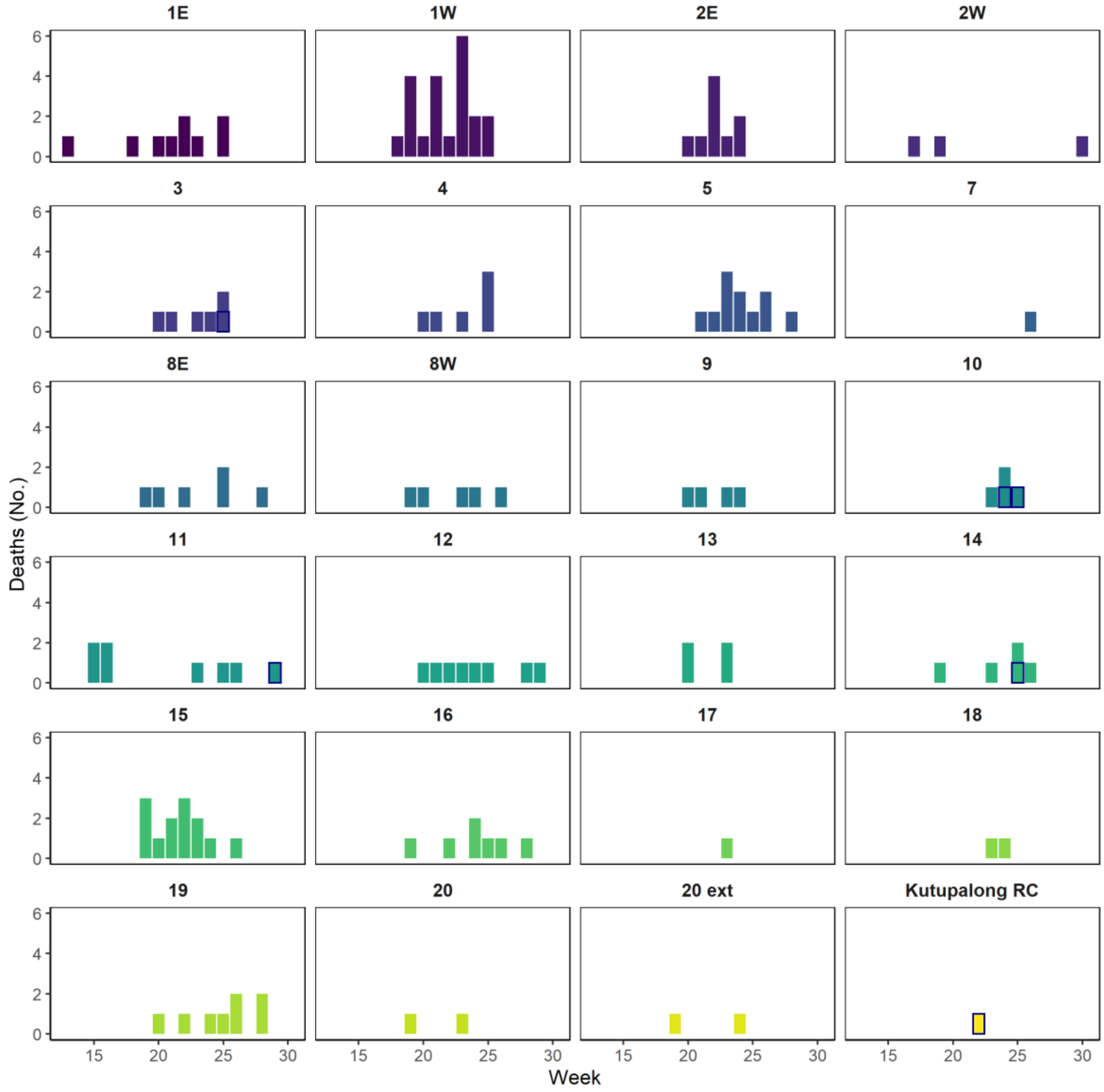


**Figure S6. Deaths by camp, weeks 13-31, 2020.** These deaths include 144 deaths identified by the IOM mortality survey and 6 deaths reported with confirmed SARS-CoV-2 infection (outlined in navy blue). In each came, identified deaths were distributed across the 19 weeks, with some camps demonstrating an outbreak-like curve (e.g., camps 1W, 2E, 9, 15). These patterns are as expected if these deaths represent true COVID-19 deaths resulting from an outbreak in this population, giving more credibility to these data.

### 3. DATA AND CODE AVAILABILITY

Aggregated, de-identified data and all code used in this analysis will be made available on https://github.com/shauntruelove/rohingyaCOVID19burden.

References


1. O'Driscoll, M. *et al.* Age-specific mortality and immunity patterns of SARS-CoV-2. *Nature* **590**, 140–145 (2021).
2. 1 in 10 in Dhaka Covid-19 positive, but mostly asymptomatic. https://bangladeshpost.net/posts/one-in-10-covid-19-patients-in-dhaka-39550.
3. Data. https://www.covid.is/data.
4. icddr,b - Press Releases. https://www.icddrb.org/quick-links/press-releases?id=97&task=view.
5. How Good are COVID-19 (SARS-CoV-2) Diagnostic PCR Tests? *College of American Pathologists* https://www.cap.org/member-resources/articles/how-good-are-covid-19-sars-cov-2-diagnostic-pcr-tests.
6. Sarker, A. R. *et al.* Prevalence and Health Care–Seeking Behavior for Childhood Diarrheal Disease in Bangladesh. *Glob. Pediatr. Health* **3**, 2333794X16680901 (2016).
7. McGowan, C. R. *et al.* COVID-19 testing acceptability and uptake amongst the Rohingya and host community in Camp 21, Teknaf, Bangladesh. *Confl. Health* **14**, 74 (2020).
8. Bhatia, A. *et al.* The Rohingya in Cox's Bazar. *Health Hum. Rights* **20**, 105–122 (2018).
9. Cai, R., Novosad, P., Tandel, V., Asher, S. & Malani, A. *Representative Estimates of COVID-19 Infection Fatality Rates from Three Locations in India*. http://medrxiv.org/lookup/doi/10.1101/2021.01.05.21249264 (2021) doi:10.1101/2021.01.05.21249264.
